# Understanding pretreatment loss to follow-up of tuberculosis patients: an explanatory qualitative study in Chennai, India

**DOI:** 10.1101/19006312

**Authors:** Beena E. Thomas, Chandra Suresh, J. Lavanya, Mika M. Lindsley, Amith T. Galivanche, Senthil Sellappan, Senthanro Ovung, Amritha Aravind, Savari Lincy, Agnes Lawrence Raja, S. Kokila, B. Javeed, S. Arumugam, Kenneth H. Mayer, Soumya Swaminathan, Ramnath Subbaraman

**Affiliations:** Department of Social and Behavioural Research, National Institute for Research in Tuberculosis, Chennai, India; District Tuberculosis Office, Chennai, India; Department of Public Health and Community Medicine and Center for Global Public Health, Tufts University School of Medicine, Boston, USA; The Fenway Institute, Fenway Health, Boston, USA; Division of Infectious Diseases, Beth Israel Deaconess Medical Center and Harvard Medical School, Boston, USA; World Health Organization Headquarters, Geneva, Switzerland; Division of Geographic Medicine and Infectious Diseases, Tufts Medical Center, Boston, USA

**Keywords:** tuberculosis, cascade of care, pretreatment loss to follow-up, initial default, implementation science, operations research, qualitative research

## Abstract

**Introduction:** Pretreatment loss to follow-up (PTLFU)—dropout of patients after diagnosis but before registration in treatment—is a major gap in TB care delivery in India and globally. Patient and healthcare worker (HCW) perspectives are critical for developing interventions to address this problem.

**Methods:** We prospectively tracked newly diagnosed smear-positive TB patients from 22 TB diagnostic centers in Chennai, one of India’s largest cities. Patients who did not start therapy within 14 days, or who died or were lost to follow-up before official registration in treatment, were classified as PTLFU cases. We conducted qualitative interviews with all trackable PTLFU patients, or family members of patients who had died. We conducted focus group discussions (FGDs) with three types of HCWs involved in TB care. Interview and FGD recordings were transcribed, coded, and analyzed with the support of Dedoose 8.0.35 software to identify key themes. We created categories into which these themes clustered, identified relationships among thematic categories, and assembled findings into a broader explanatory model for PTLFU.

**Results:** We conducted six FGDs comprising 53 HCWs and 33 individual patient or family member interviews. Themes clustered into five categories. Examining relationships among these categories revealed two pathways leading to PTLFU as part of a broader explanatory model. In the first pathway, administrative and organizational health system barriers—including the complexity of navigating the system, healthcare worker absenteeism, and infrastructure failures—resulted in patients feeling frustration or resignation, leading to disengagement from care. Health system barriers experienced by patients were in turn shaped by constraints that made it difficult for HCWs to do their jobs effectively. In the second pathway, negative or judgmental HCW attitudes and behaviors towards patients contributed to patient distrust of the health system, resulting in refusal of further care.

**Conclusion:** Health system barriers contribute substantially to PTLFU directly and by amplifying patient-related challenges to engaging in care. Interventions should focus on removing administrative hurdles patients face in the health system, improving the quality of the HCW-patient interaction, and alleviating constraints HCWs face in being able to provide optimal patient-centered care.

## Introduction

Tuberculosis (TB) care delivery may be undermined by patient loss to follow-up across sequential steps from care-seeking to diagnosis to successful treatment, comprising the cascade of care.^1^ Pretreatment loss to follow-up (PTLFU), which refers to patient losses after TB diagnosis but before registration in treatment, is a key gap in the care cascade in several high TB burden countries.^2 3^ An analysis conducted in India, which accounts for one-fourth of TB cases and one-third of TB deaths globally, estimated that >200,000 patients experience PTLFU annually in the national TB program.^4^

Prior studies of PTLFU in India have tracked down “lost” patients to obtain their perspectives on why they dropped out of care. While these studies provide valuable information, they have generally reported a list of reasons patients did not start TB treatment, without providing in-depth analysis of their narrative responses or producing a framework that can inform actions to address this problem. Most of these studies assumed that PTLFU results from a decision by the TB patient not to seek further care, rather than from a more complex interaction between the patient and the health system. As such, reasons for PTLFU described in these studies have generally focused on patient factors, including presentation with advanced illness,^5 6^ low literacy,^7^ employment-related barriers,^6 8 9^ alcohol use,^8^ denial of diagnosis,^7^ urban-rural movement,^6-11^ and not collecting sputum test results.^5 7^ Health system barriers contributing to PTLFU often emerged indirectly in these studies, including patient dissatisfaction with health services,^5-8^ patient concerns about being monitored via facility-based directly observed therapy (DOT),^5 7^ or inability of healthcare workers (HCWs) to find patients due to poor recording of contact information.^6 7 12^

In this paper, we analyze qualitative data from TB patients and HCWs with the goal of proposing an explanatory model for PTLFU that can inform future interventions. Our analytical approach is similar to those used in recent papers that integrated thematic findings across single or multiple studies to propose explanatory models for non-initiation of HIV treatment,^13^ disengagement from HIV care,^14^ or non-adherence to HIV or TB medications.^15-17^ Qualitative data collection was embedded within a recent cohort study in the government TB program in Chennai, one of India’s largest cities, which found that more than one-fifth of smear-positive TB patients experienced PTLFU.^18^ The cohort study also showed that patients with a prior TB treatment history were at higher risk for PTLFU, which is concerning because these patients are more likely to have drug-resistant TB, such that failure to start treatment could contribute to transmission of drug-resistant strains.

Although this previous quantitative study identified *who* is more likely to experience PTLFU, it did not shed light on *why* these patients with a life-threatening disease did not officially register in treatment. Our qualitative analysis in the current manuscript provides insights into the problem of PTLFU that cannot be gleaned using quantitative methods alone. We perform a thematic synthesis using the qualitative data and create an explanatory model that can inform interventions to reduce PTLFU in urban India.

## Methods

### Study setting

Chennai has a population of 7.1 million people and a high TB prevalence of about 349 per 100,000 people.^19^ The city’s TB services follow guidelines of India’s Revised National TB Control Programme (RNTCP). The current study followed newly diagnosed TB patients at 22 of Chennai’s RNTCP-designated microscopy centers (DMCs), which accounted for 90% of all smear-positive TB patients diagnosed in the city’s government sector in 2014.^18 20^ Four of these DMCs, located in specialized TB facilities or tertiary hospitals, diagnosed more than half of the city’s smear-positive patients;^20^ we refer to these four as “high-volume DMCs” and to the remaining 18 as “moderate- or low-volume DMCs.”

### Case definitions for pretreatment loss to follow-up

We discuss PTLFU case definitions briefly, as they have been described in detail in the prior quantitative manuscript.^18^ Patients diagnosed with smear-positive TB had to traverse a multistep process of diagnosis and linkage to care, which differed for those initially managed as outpatients or inpatients. For both types of patients, we identified three stages at which patients were lost (Fig. 1): during the diagnostic workup; during outpatient referral or hospital admission; or during the official RNTCP registration process, because registration was delayed for more than two weeks for nearly one-third of patients.^18^ Patients who completed all three stages to get officially registered were considered to have successfully linked to care. Conversely, we defined PTLFU as including two types of patients: (1) those who did not start TB therapy at a government DOT center within 14 days of initial sputum evaluation, due to loss to follow-up, death, or delay; or (2) those who started TB therapy but were lost to follow-up or died before official registration.^18^ For this qualitative study, our PTLFU patient sample therefore included individuals who dropped out at different points in this multistep process, providing insights into the diverse challenges patients face in linking to care.

**Fig. 1.**
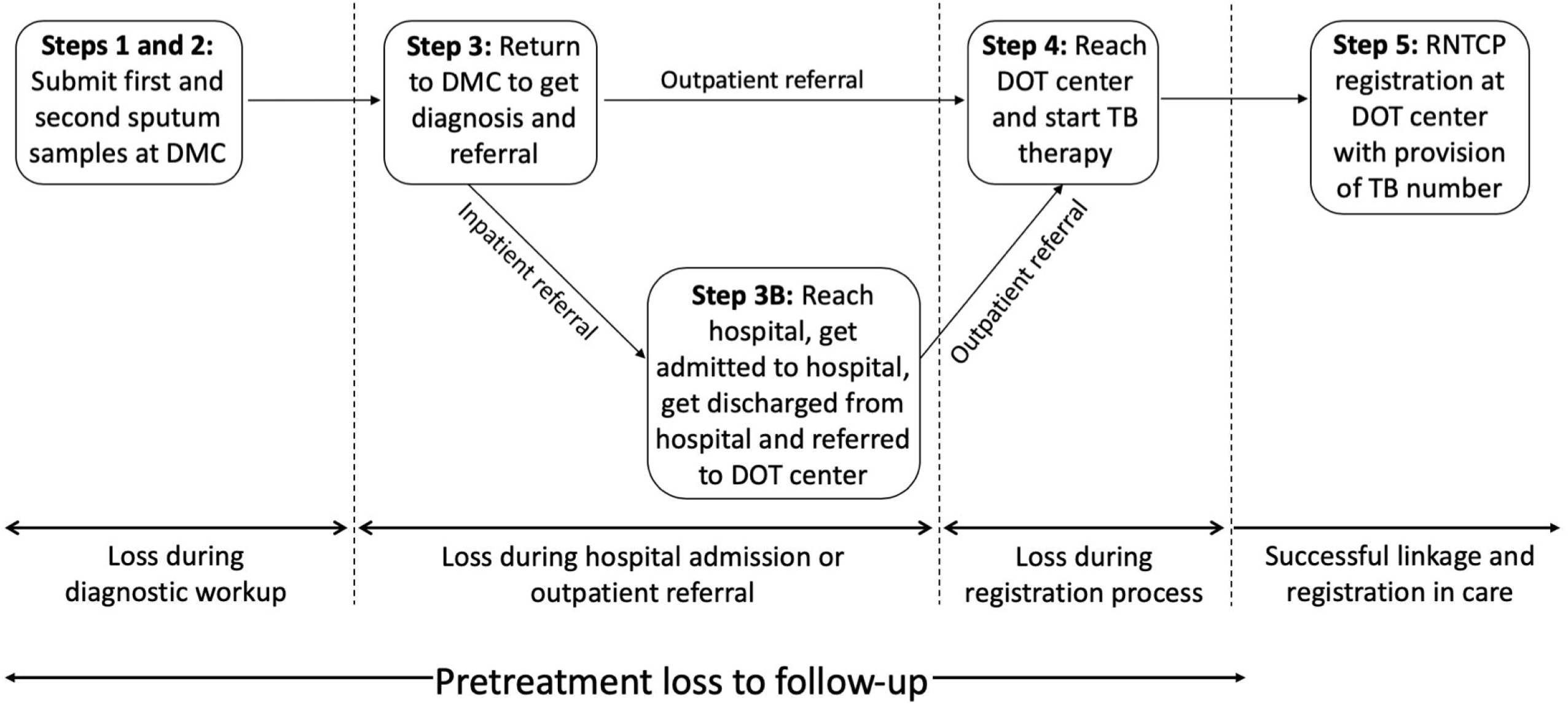
Multistep process of diagnosis and linkage to care for smear-positive tuberculosis patients in Chennai, India. Modified from Thomas et al. 2018.^18^ DOT, directly observed therapy; DMC, designated microscopy center; TB, tuberculosis.

### Field methods for evaluating pretreatment loss to follow-up

Between October 2015 and June 2016, we followed all smear-positive TB patients ≥18 years old who were diagnosed during a four-week time period at each of the 22 DMCs.^18^ Patients were tracked starting no earlier than 14 and no later than 21 days after their first positive sputum test, using a systematic process involving audits of health records, phone outreach, and home visits to patients if necessary.^18^ We continued to follow patients who started treatment within 14 days until they were officially registered with provision of a TB Number. We classified patients as being untrackable if researchers were unable to find them after multiple phone calls and at least one home visit.

### Collection of qualitative patient data

We collected qualitative interviews between October 2015 and June 2016, concordant with patient tracking efforts for the cohort study. Interviews lasted 30 to 45 minutes and were conducted at a location of the patient’s (or her family’s) choosing by researchers with a master’s degree in social work. After obtaining informed consent, interviews were audio-recorded in the Tamil language and translated later to produce English-language transcripts.

Out of 76 PTLFU patients in the cohort study, 27 were untrackable and could not be interviewed.^18^ Six PTLFU patients declined to be interviewed, due to denial of their TB diagnosis (two patients) or unclear reasons (four patients). The audio recording for one interview was muffled and could not be analyzed. Nineteen PTLFU patients died after starting TB therapy but before official registration. We interviewed 10 of these patients, because thematic saturation was achieved for this sub-group based on coding of these initial transcripts. As such, our findings are reported for n=33 PTLFU patients.

Interview guides were used to elicit narrative information on the timeline of events and challenges that led to PTLFU. Questions varied based on the time at which a patient was “lost” (i.e., during diagnostic workup, outpatient referral, hospital admission, or registration process). For example, for patients who did not reach the DOT center to start treatment, a key open-ended question was “What were reasons you were not able to make it to the [DOT] center to start treatment?” Language was modified as needed for interviews conducted with patients’ family members.

### Collection of qualitative healthcare worker data

Between July and December 2016, we conducted six HCW FGDs. Two FGDs each were conducted with three types of auxiliary health personnel in Chennai’s TB program: (1) Laboratory Technicians (LTs), who collect sputum samples, record patients’ contact information, and evaluate sputum samples for evidence of acid-fast bacilli; (2) Health Visitors (HVs), who counsel and refer patients (at DMCs) or start them on TB treatment (at DOT centers); and (3) Senior Treatment Supervisors (STSs), who supervise HVs at multiple DOT centers.

The six FGDs were conducted after monthly meetings of HCWs from across the city at Chennai’s district TB office. As such, the FGDs included a relatively representative sample of HCWs from geographically diverse areas of the city and all levels of health facilities (i.e., primary, secondary and tertiary centers). FGDs lasted 45 to 60 minutes and were conducted in Tamil by researchers with social work backgrounds. FGD guides included open-ended questions to elicit group feedback on factors contributing to PTLFU and problems identified in the quantitative study, such as delayed registration of patients.^18^

### Qualitative data analysis

We used an inductive approaches for the thematic analysis of interviews and FGDs.^21 22^ We created an initial coding scheme based on discussion within the research team, including researchers who collected the interviews and FGDs. Interview and FGD transcripts were independently coded by three researchers using Dedoose software (version 8.0.35, Los Angeles, CA: SocioCultural Research Consultants, LLC; www.dedoose.com). Researchers met frequently to reconcile differences in application of codes and to identify new themes emerging from the data. After incorporating new themes into the coding scheme, all interviews were coded a second time.

We employ concepts from meta-ethnography^23^ and thematic synthesis,^24^ approaches that have previously been used to analyze themes across single or multiple studies and develop explanatory models for non-engagement of patients in HIV or TB care.^13-16^ Using Dedoose, we identified common or salient themes (i.e., codes) to produce a list of unique reasons for why PTLFU occurs. We extracted representative quotations for these themes and reviewed these data again to create thematic categories (i.e., a cluster of themes representing a broader construct). We examined representative quotations within categories to map out relationships among these categories and assemble a broader explanatory model for PTLFU.

### Patient and Public Involvement

This study protocol was reviewed, with critical feedback provided, by the Community Advisory Board of the National Institute for Research in TB in Chennai, which includes public stakeholders in the community as well as former TB patients and patient advocates.

## Results

### Descriptive characteristics of study participants

Of the 33 in-depth TB patient interviews, 13 (39%) were conducted with family members of patients who had died. The patients’ ages ranged from 21 to 80 (median 53) years. Of the sample, 3 (9%) were women; 4 (12%) lived outside of Chennai in surrounding rural areas; and 25 (76%) were diagnosed at a high-volume DMC (i.e., tertiary hospital or specialized TB facility). Notably, 21 (64%) had a prior TB treatment history, which included those who completed their prior TB treatment course and those who were lost to follow-up before finishing treatment.

The six FGDs included 53 participants, of whom 18 (34%) were STSs, 18 (34%) were HVs, and 17 (32%) were LTs. Sex and years of work experience in the RNTCP were captured for 40 participants (i.e., four of the FGDs), of whom 30 (75%) were men. Years of work experience ranged from 1 to 30 (median 5.5).

### Thematic categories

Barriers contributing to PTLFU clustered into five thematic categories. Three categories reflected TB patient experiences, attitudes, or behaviors, and these categories primarily emerged from the in-depth interviews with patients or their family members: (1) patient-, family-, and society-related challenges contributing to PLTFU that are largely independent of the health system; (2) health system organizational and administrative barriers experienced by patients; and (3) patient attitudes and behaviors towards the health system that contribute to PTLFU. The remaining two thematic categories reflected HCW experiences, attitudes, or behaviors, and these findings primarily emerged from the FGDs: (4) social and resource constraints in the health system that limit HCWs’ ability to do their jobs effectively; and (5) HCW attitudes and behaviors towards patients that contribute to PTLFU.

### Thematic category 1. Patient-, family-, and society-related challenges that are largely independent of the health system

Some themes contributing to PTLFU reflected challenges faced by patients at the individual level or in relation to broader society that were not related to interactions with the health system (Table 1). At the individual level, presentation with advanced illness from TB (Q1) and comorbidities such as alcohol use disorder (Q2) and depression made it difficult for patients to navigate health facilities and contributed to deaths prior to official registration. At the level of patients’ families, some patients, especially women, lacked social support to be accompanied to medical visits (Q3), experienced TB-related stigma (Q3), or did not follow-up with sputum test results because they were attending family functions or festivals, often in villages outside of Chennai (Q4). At the societal level, employment (Q5), poverty (Q6), homelessness, and weather-related constraints contributed to PTLFU.

**Table 1.** Representative quotations regarding patient-, family-, and society-related challenges

| Challenge | Representative quotation |
| --- | --- |
| Presentation with advanced illness | Q1. *Even at the time of admission [to the hospital], the doctor said that his illness was too advanced. So the doctor advised me to take care of him at home, but I refused . . . Three days after talking to the doctor my father died.* (Family member of a 34-year-old man with a prior treatment history). |
| Alcohol use disorder | Q2. *He consumed alcohol. For that reason, he didn't take TB medications* |
|  | *properly.* (Family member of a 26-year-old man with a prior treatment history). |
| Lack of social support and enacted stigma | Q3. *I came here without my family members' knowledge, as they otherwise would not allow me to visit [this specialty hospital]. If others learned that I am visiting [this hospital], they will tag me as being a TB patient.* (75-year-old woman without a prior treatment history) |
| Attending family function or festival | Q4. *I went back to my native village for [my grandfather's] funeral rites. I had to take care of all formalities, such as paying for funeral expenses. Due to this family situation, I couldn't go back to pick up my TB test report on time.* (31-year-old man without a prior treatment history) |
| Work constraints | Q5. *I got my [sputum test] report only after one week due to work. It was very difficult to get leave approval from my office, and my family depends on my salary, as I am getting daily wages.* (60-year-old man with a prior treatment history). |
| Poverty | Q6. *I did not bring my brother immediately [to the hospital] because we don't have the money.* (Family member of a 35-year-old man with a prior treatment history) |

### Thematic category 2. Organizational and administrative barriers in the health system experienced by patients

Thematic category 2 emerged as the most prominent set of problems contributing to PTLFU (Table 2). These barriers reflected organizational and administrative barriers within that made it challenging for patients to navigate the health system. Patients described difficulties finding different outpatient clinics within tertiary hospitals (Q7), having to return to the same health facility repeatedly, and having to navigate between facilities for diagnostic workup and treatment initiation (Q8). Inadequate communication of information by HCWs contributed to navigational problems (Q8) and other problems, such as patients not being informed of their TB diagnoses or having low motivation to pursue next steps in care because of lack of general information about TB (Q9).

**Table 2.** Representative quotations regarding organizational and administrative barriers in the health system experienced by patients

| Health system barrier | Representative quotation |
| --- | --- |
| Complexity of navigation within facilities | Q7. *We went to [a large tertiary hospital] for initial checkup . . . They didn’t tell us much. They said go to number 3 [outpatient clinic] and then number 5 [outpatient clinic] for two days. After running from pillar to post, we just gave up and returned home.* (Family member of a 80-year-old man without a prior treatment history who had died). |
| Complexity of navigation between facilities | Q8. *[The HCW] told me to go to Old General Hospital. I thought that by “Old General Hospital” she meant Madras Medical College, which is very far away from where I live. So I went back home . . . I later found out that Old General Hospital meant [a different local hospital], which is nearby, just opposite this clinic.* (58-year-old man without a prior treatment history). |
| Failure to notify patient of TB diagnosis or to provide general information about TB | Q9. *They referred me to [a] hospital for treatment . . . But they did not say anything about my health condition and about my disease.* (45-year-old man without a prior treatment history). |
| Infrastructure failures | Q10. *Thrice I came to [a tertiary hospital] to receive my test report but . . . [t]hey said, ‘ . . . [Y]ou have to wait for some days to collect your report. We can prepare your report only when the power supply is available.’* (75-year- |
|  | old woman without a prior treatment history). |
| Healthcare worker demanded documents | Q11. *[After the patient reached the rural DOT center, the HCW said]: ‘Go back to [tertiary hospital where the patient was diagnosed in the city] and bring a referral slip—only then can we start treatment.’* (72-year-old man without a prior treatment history). |
| Healthcare worker absenteeism | Q12. *After diagnosis, they referred me to [a local primary health center] . . . the HCW referred me from there to [a second primary health center] . . . The next day I went to that [second primary health center] . . . However, at that time, the hospital staff asked me to return the next day, because the responsible healthcare worker was not available.* (40-year-old man with a prior treatment history). |
|  | Q13. *The X-ray technician said that he . . . can only give the result tomorrow. I pleaded with him. I told him I felt too ill. He asked me to come after one hour . . . At that time the doctor had left the hospital. The Sister [nurse] asked me to come back on Wednesday.* (43-year-old man with a prior treatment history). |
| Rigidity of facility-based DOT | Q14. *[HCWs who perform DOT] come at 11 am. Patients will be standing in the queue; they call us one by one to give the tablets. We need to take tablets in front of them and sign. It almost takes until 12 pm. Then how can I go to work?* (43-year-old man with a prior treatment history). |

Even when they navigated to the right location, patients often ran into functional or administrative hurdles that impeded further care, such as infrastructure failures (e.g., of computers or electricity, Q10) or demands by HCWs to present referral documents from sites where patients were initially diagnosed (Q11). HCW absenteeism was a hurdle to patients progressing along the care pathway (Q12, Q13). Prior experiences with facility-based DOT—the main treatment monitoring approach at the time of this study—deterred some patients with a prior TB treatment history, who had found multiple visits each week to DOT centers to be incompatible with work and other life demands (Q14).

These health system barriers often intersected. For example, navigational challenges were often complicated by healthcare worker absenteeism. As such, the effect of these hurdles was often cumulative, eventually provoking feelings of frustration or resignation that led patients to passively disengage from the process of linkage to care—as opposed to actively refusing care. One patient’s family characteristically described this sense of resignation as follows: “*After running from pillar to post, we just gave up and returned home*.” (Q7).

### Thematic category 3. Negative patient attitudes and behaviors towards the health system

Some patients or their family members expressed a different set of emotions, attitudes, and behaviors that emerged in response to prior or current interactions with the health system (Table 3). These included distrust of the health system (Q15, Q16) and fear of medical care (Q17), including fear of medication toxicities (Q18). Such attitudes were more common in patients with a prior TB treatment history and reflected previous negative experiences with the government TB program.

**Table 3.** Representative quotations regarding patient attitudes and behaviors towards the health system

| Patient attitude or behavior | Representative quotation |
| --- | --- |
| Distrust of the health system | Q15. *I know about [the nearby government hospital]. I don't like that place. So I came to [a tertiary hospital]. In [the tertiary hospital], they maintain cleanliness.* (60-year-old man with a prior treatment history). |
|  | Q16. *The doctor was asking the Sister (nurse) whether he could give me the tablet or not. I got worried when I heard the doctor asking the nurse whether he should be prescribing a particular tablet.* (43-year old man with a prior treatment history). |
| Fear of further medical care | Q17. *Yes, the doctor admitted him in the ward . . . at that time, two patients died close to my husband . . . the next night two more patients also died there so my husband became very scared and we discharged him.* (Family member of an 80-year-old man without a prior treatment history). |
| Fear of medication toxicities | Q18. *He would tell me, 'They are going to kill me by giving me these tablets. If I take these tablets, I will feel very giddy.'* (Family member of a 45-year-old man with a prior treatment history) |
| Care-seeking at multiple sites | Q19. *I didn't get proper treatment at [the first tertiary care hospital] due to lack of staff at the facility, and I left the [second tertiary hospital] due to lack of hygiene and cleanliness. So I decided to go to [a third facility] for further care.* (55-year-old man without a prior treatment history). |
| Refusal of further care | Q20. *When they gave him tablets, he would keep them in his hands and throw them away later. That is what happened at [the inpatient tertiary care hospital] also. They gave him tablets, and he used to go to the bathroom and throw them away. After few days, he ran away from the hospital.* (Family member of a 57-year-old man with a prior treatment history). |

These attitudes negatively affected patients’ care-seeking behavior. While patients were often referred from one health facility to another by HCWs, in some cases, patients themselves sought care at multiple health facilities, without being referred (Q19). Care seeking at multiple sites resulted in patients not following up on sputum microscopy results or in delays in reaching treatment centers. The term “doctor shopping” is sometimes used to describe this behavior in India; however, this term implies a degree of patient choice. In contrast, patients usually sought care at multiple sites because of concerns about quality of care or hygiene at previous facilities.

While patients responded to organizational and administrative health system barriers by passively disengaging from care, attitudes and behaviors that reflected distrust of the health system usually resulted in active refusal of further care (Q17, Q18, and Q20).

### Thematic category 4. Social and resource constraints in the health system that limit the ability of HCWs to effectively do their jobs

HCWs reported numerous constraints that limited their ability to effectively perform their jobs (Table 4). All three types of HCWs reported having to work at multiple health facilities (Q21), which was particularly problematic for those who were supposed to interact with patients on a daily basis during diagnostic workup (LTs) or treatment initiation (HVs). These HCWs could not follow-up with patients as required, since they had to be working at other sites. Absence of RNTCP staff also contributed to poor recording of patient contact information, because non-RNTCP staff in health facilities were not sure how to correctly record this information or did not understand its importance (Q22). At high-volume facilities, HCWs had little time to spend accurately recording patient contact information (Q23).

**Table 4.**
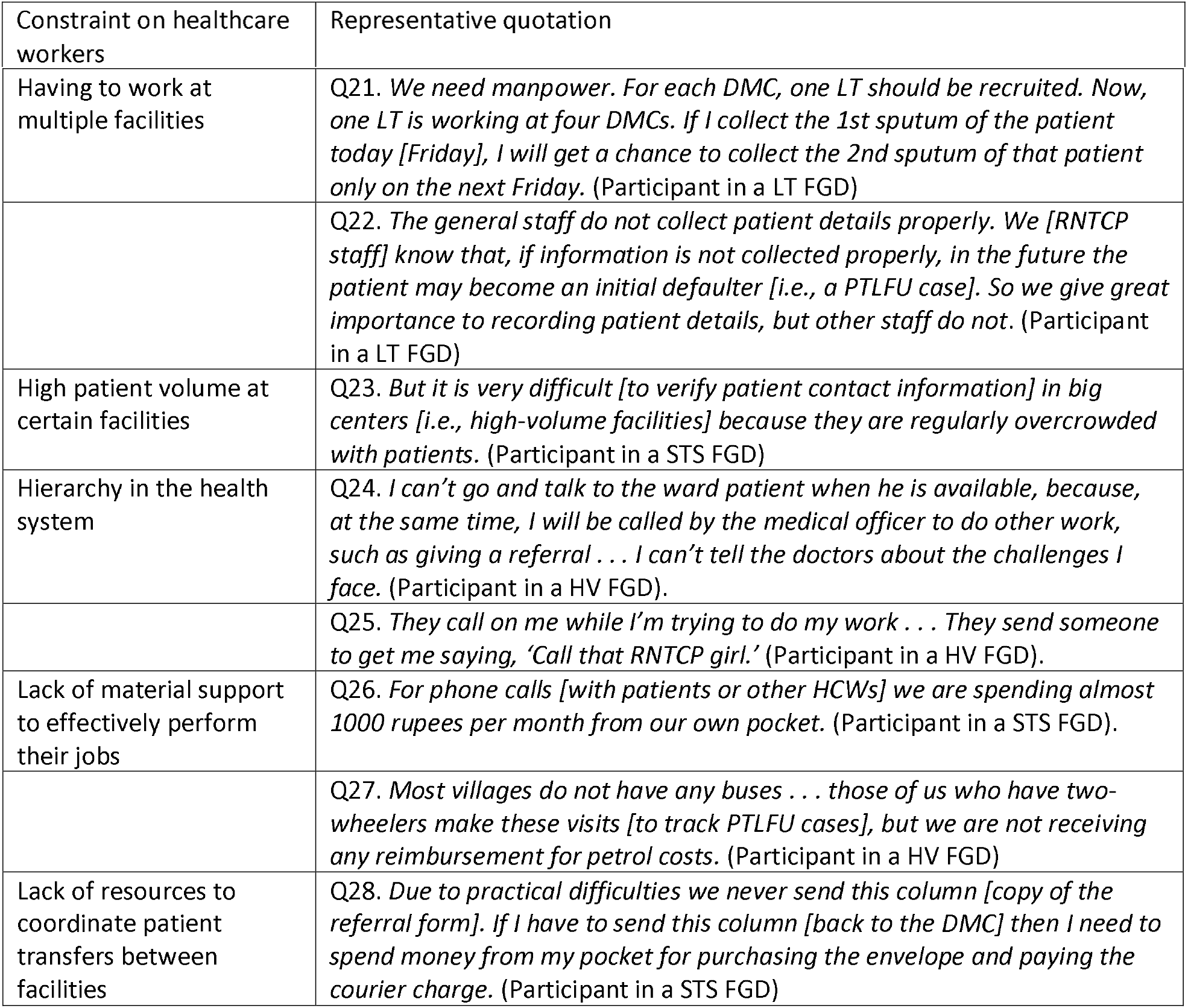
Representative quotations regarding social and resource constraints in the health system that limit the ability of HCWs to effectively do their jobs

| Constraint on healthcare workers | Representative quotation |
| --- | --- |
| Having to work at multiple facilities | Q21. *We need manpower. For each DMC, one LT should be recruited. Now, one LT is working at four DMCs. If I collect the 1st sputum of the patient today [Friday], I will get a chance to collect the 2nd sputum of that patient only on the next Friday.* (Participant in a LT FGD) |
|  | Q22. *The general staff do not collect patient details properly. We [RNTCP staff] know that, if information is not collected properly, in the future the patient may become an initial defaulter [i.e., a PTLFU case]. So we give great importance to recording patient details, but other staff do not.* (Participant in a LT FGD) |
| High patient volume at certain facilities | Q23. *But it is very difficult [to verify patient contact information] in big centers [i.e., high-volume facilities] because they are regularly overcrowded with patients.* (Participant in a STS FGD) |
| Hierarchy in the health system | Q24. *I can't go and talk to the ward patient when he is available, because, at the same time, I will be called by the medical officer to do other work, such as giving a referral . . . I can't tell the doctors about the challenges I face.* (Participant in a HV FGD). |
|  | Q25. *They call on me while I'm trying to do my work . . . They send someone to get me saying, 'Call that RNTCP girl.'* (Participant in a HV FGD). |
| Lack of material support to effectively perform their jobs | Q26. *For phone calls [with patients or other HCWs] we are spending almost 1000 rupees per month from our own pocket.* (Participant in a STS FGD). |
|  | Q27. *Most villages do not have any buses . . . those of us who have two-wheelers make these visits [to track PTLFU cases], but we are not receiving any reimbursement for petrol costs.* (Participant in a HV FGD) |
| Lack of resources to coordinate patient transfers between facilities | Q28. *Due to practical difficulties we never send this column [copy of the referral form]. If I have to send this column [back to the DMC] then I need to spend money from my pocket for purchasing the envelope and paying the courier charge.* (Participant in a STS FGD) |

HCWs were unable to complete routine tasks, including patient counseling, because of hierarchy in the health system, since LTs, HVs, and STSs fall under the supervision of doctors and nurses (Q24, Q25). Supervisors pulled HCWs away from data entry tasks and interactions with TB patients; HCWs were ordered to perform tasks unrelated to their primary job taking care of TB patients. HCWs lacked material support (e.g., reimbursement of costs) to effectively perform their jobs, including communicating with patients or other HCWs by phone (Q22) and tracking patients whose homes were inaccessible by public transportation (Q27). Resource constraints hindered coordination between facilities when patients were referred, since HCWs did not have money for phone or mail communication (Q28).

### Thematic category 5. Negative HCW attitudes and behaviors towards patients

Examples of negative or judgmental HCW attitudes and behaviors towards patients emerged in both HCW FGDs and patient interviews (Table 5). HCWs scolded patients (Q29), especially those with a prior TB treatment history. Some patients and HCWs reported HCW behaviors that were perceived to stigmatize patients (Q30). HCWs would sometimes notify community leaders or members of self-help groups (for female patients) about a patient’s TB diagnosis with the goal of encouraging social support; however, HCWs described how such actions sometimes irreparably harmed their relationships with patients, who perceived these behaviors as violating their autonomy (Q31). Coercive behavior aimed at encouraging treatment adherence sometimes resulted in poorer engagement in care by patients (Q32). Patients described other negative HCW behaviors, such as asking for bribes (Q33). At its most extreme, this spectrum of HCW behaviors included delaying registration (Q34) or refusing care to patients perceived to be “irregular” or “disobedient” (Q35).

**Table 5.** Representative quotations regarding negative HCW attitudes and behaviors towards patients

| HCW attitude or behavior | Representative quotation |
| --- | --- |
| Scolding patients | Q29. *I told [the doctor] that my health condition improved quickly during my first TB treatment, which is why I had stopped therapy early. He said, ‘Because of your disobedience in following your last treatment instructions, you are now suffering again.’* (60-year-old man with a history of prior treatment). |
| Stigmatizing behavior | Q30. *Patients believe the doctors more than us, but the doctors don’t have enough time to talk to patients. The staff nurse can counsel the patients, but they treat the patients as untouchables.* (Participant in a HV FGD). |
| Violation of confidentiality or disclosure of diagnosis | Q31. *Patients will ask with us, ‘Why did you share my information with the panchayat [community] leader? . . . Who gave you the right to share information regarding my disease condition with others? . . . What will others think about me?’* (Participant in a HV FGD). |
| Coercion | Q32. *Sometime we used to blackmail the irregular patients by using these permanent ID proof. For example, we got a driving license as a proof of identification from an auto driver [patient]. Then we said to him, ‘If you take treatment irregularly, we will cancel your driving license.’* (Participant in a STS FGD). |
| Demanding bribes for further care | Q33. *Ward staff in the hospital were asking for bribes for towing patients to the X-ray department.* (40-year-old man with a prior treatment history). |
| Delays in patient registration | Q34. *We generally assign TB Numbers within a week or within two weeks. If the patient is regular [on TB therapy], then we assign the TB Number the same day, but in the case of category II patients [i.e., those with a prior TB treatment history], we take two or more days for giving the TB Number.* (Participant in a STS FGD). |
| Refusal of care to patients | Q35. *They [nurses or supervising providers] do not want to care for the patient. For default cases [i.e., those with a prior TB treatment history] . . . the nurse will say, ‘Send him out. Don’t give him a strip of tablet.’ Or, ‘Why are you giving him tablets?’* (Participant in a HV FGD). |

### Interconnections among thematic categories: toward an explanatory model

Upon further evaluation, we identified a series of interconnections among four of the thematic categories (2, 3, 4, and 5) that suggest a broader explanatory model. Our model only focuses on the health system’s contribution to PTLFU and, for that reason, does not integrate findings from thematic category 1, which described patient-, family-, and society-related challenges that are largely independent of the health system. Health system barriers may serve to amplify challenges in thematic category 1, however. In the following section, we provide specific examples, using linked representative quotations, to justify how thematic categories connect to reveal these two different pathways by which patients’ interactions with the health system lead to PTLFU (Fig. 2). In addition, there may be logical interconnections between the two pathways (dotted lines in Fig. 2), which we do not discuss in detail.

**Fig 2.**
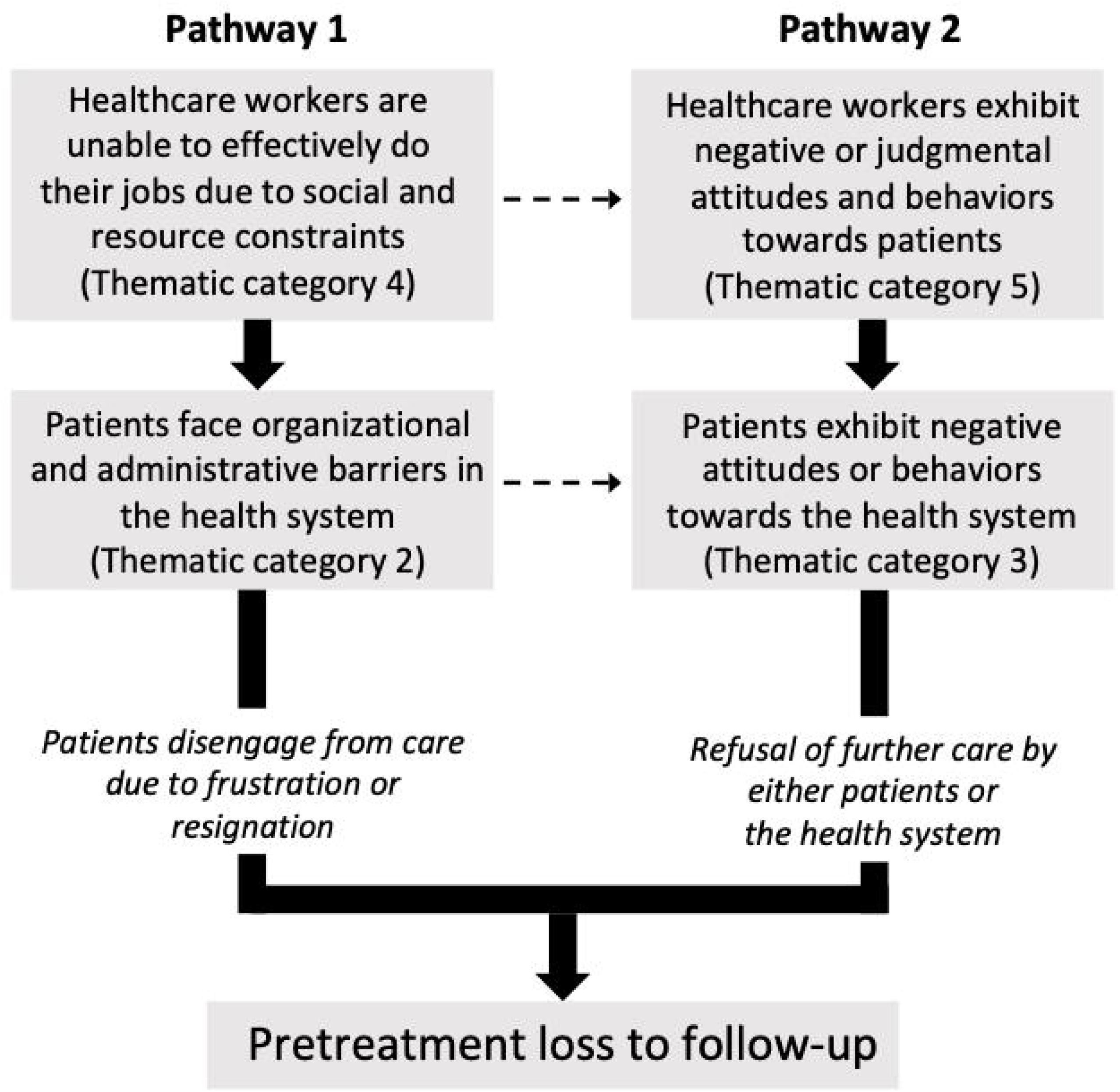
An explanatory model for the health system’s role in contributing to pretreatment loss to follow-up (PTLFU) of tuberculosis (TB) patients in Chennai, India. Dark arrows represent the main pathways contributing to PTLFU. Smaller dotted arrows represent possible connections between these pathways.

### Pathway 1

In the first pathway, constraints faced by HCWs (thematic category 4) directly contributed to organizational and administrative health system barriers (thematic category 2). Most patients in this pathway wanted to seek further TB care but passively disengaged from the care pathway due to frustration or resignation.

For example, some patients showed up to health facilities for the next step in care (e.g., picking up a test result, starting therapy), only to find that the relevant HCW was not available. From the HCW perspective, the reason for this problem was clear: all three types of HCWs were responsible for providing care at multiple health facilities. For example, HVs described the following challenges:

*HV 1: We are covering more than one [health facility] madam…*

*HV 1: Four centers madam*.

*HV 2: Two centers*.

*HV 6: Six centers madam*.

*HV 9: For this reason, we aren’t able to treat patients well, madam*.

(Excerpt from a HV FGD)

LTs, who are responsible for recording patient contact information and evaluating sputum samples, described similar challenges:

*We need manpower … Right now, one LT is working at four DMCs. If I collect the 1st sputum of the patient today [Friday], I will get a chance to collect the 2nd sputum of that patient only on the next Friday*.

(Participant in a LT FGD)

Patients experience the problem of HCWs working at multiple facilities as HCW absenteeism:

*After diagnosis, they referred me to [a local primary health center] … the HCW referred me from there to [a second primary health center]* … *The next day I went to that [second primary health center] … However, at that time, the hospital staff asked me to return the next day, because the responsible HCW was not available*.

(40-year-old man with a prior treatment history).

*The X-ray technician said that he … can only give the result tomorrow. I pleaded with him. I told him I felt too ill. He asked me to come after one hour … At that time the doctor had left the hospital. The Sister [nurse] asked me to come back on Wednesday*.

(43-year-old man with a prior treatment history).

As another example, poorly recorded patient contact information was a factor contributing to PTLFU in our prior quantitative study.^18^ LTs and STSs at high-volume centers reported having little time to record and verify contact information:

*But it is very difficult [to verify patient contact information] in big centers [i*.*e*., *high-volume facilities] because they are regularly overcrowded with patients*.

(Participant in a STS FGD)

This problem of poorly recorded contact information was experienced by patients as lack of outreach by the health system, even for critical information such as the patient’s diagnosis:

*No one told me about my TB diagnosis*.

(62-year-old woman without a prior treatment history).

While we will not present them in detail here, we identified other examples of how constraints on HCWs contributed to health system barriers faced by patients, leading to disengagement from care. For example, in the context of health system hierarchy, supervisors pulled HCWs away from their interactions with patients, lowering patients’ motivation, since they were not given adequate information about TB. As another example, due to lack of monetary support, HCWs at DMCs were hesitant to spend out-of-pocket for phone and postal fees to notify DOT centers of patient referrals or for transportation fees to visit patients’ homes, which exacerbated patients’ navigational challenges.

### Pathway 2

In the second pathway, negative or judgmental HCWs attitudes and behaviors towards patients (thematic category 5) shape negative patient attitudes and behaviors towards the health system (thematic category 3), leading to refusal of further care by either the patient or the health system (Fig. 2). Active refusal of further care by patients in this pathway is in contrast to the first pathway, in which patients passively disengaged from care due to frustration or resignation.

The most common negative HCW behavior was scolding, which reflected judgmental attitudes towards perceived moral failures of patients they deemed to be “disobedient” or “irregular” with therapy. For example, one patient described the following interaction:

*I told [the doctor] that my health condition improved quickly during my first TB treatment, which is why I had stopped therapy early. He said, ‘Because of your disobedience in following your last treatment instructions, you are now suffering again*.*’*

(60-year-old man with a history of prior treatment).

Patients noted that scolding made them hesitant to pursue next steps in care:

*Some of [the HCWs] talked nicely to me, but the doctor scolded me, so I didn’t return to pick up my test result*.

(62-year-old woman without a prior treatment history).

Stigmatizing behaviors by HCWs similarly resulted in distrustful patient attitudes towards the health system. In other cases, HCWs conducted home visits or involved community members in patients’ care, such as local leaders or women’s self help group members. Such involvement was often initiated without seeking a patient’s permission, reflecting HCW attitudes that were well-intentioned but not respectful of a patient’s privacy. Patients viewed disclosure of their TB diagnoses to be a violation of autonomy.

HCWs themselves described how patients sometimes refused further care because of perceived violations of autonomy:

*We once visited one of the [PTLFU] patients with our team, including the doctor, STS, STLS, and HV. But he said ‘I feel ashamed because of your action, so I cannot take medicines*.*’*

(Participant in a HV FGD)

In the second pathway, the health system itself sometimes delayed or refused care to some patients. In many cases, these were patients with a prior TB treatment history who were presumed to be at higher risk for poor outcomes. In some cases, HCWs did not directly refuse care but instead gave these patients a trial period, in which they delayed registering these patients to see if they would return to the clinic regularly:

*STS 5: Normally in the initial days [of therapy] the patients take treatment with full enthusiasm, but later on some patients stop coming back, so we keep their treatment cards separately*.

*STS 4: In the meantime we inquire with the patient’s caretaker … even after such inquiries, a few patients don’t return, so we can’t do anything more [to retrieve them] …*

*Moderator: So you never register that case?*

*STS 5: Yes, we never register that case*.

(Excerpt from a STS FGD)

Patients were sometimes aware they had not been formally registered, which created a sense that they were being devalued by the health system:

*They opened a treatment card for me, but they didn’t give me a TB Number because they said I had been taking treatment irregularly*.

(40-year-old man with a prior treatment history)

Less commonly, HCWs directly refused care to patients perceived to be high-risk:

*They [nurses or supervising providers] do not want to care for the patient. For default cases [i*.*e*., *those with a prior TB treatment history] …the nurse will say, ‘Send him out. Don’t give him a strip of tablet*.*’*

*Or, ‘Why are you giving him tablets?’*

(Participant in a HV FGD).

Examples of situations where care was refused to patients also emerged in the patient interviews:

*I went [to the treatment center] but the Sister [nurse] said, ‘Come back tomorrow*.*’ She refused to provide me with any tablets so I became very vexed. I thought, ‘Why did I come all the way here?’ So I did not continue treatment*.

(55-year-old man with a prior treatment history).

## Discussion

In this qualitative study of TB patients and HCWs in one of India’s largest cities, our explanatory model highlights the critical role of the health system in contributing to PTLFU. We identified two health system pathways leading to poor outcomes, each of which has distinct implications for developing future interventions to address this gap in care. In the first pathway, patients faced various organizational and administrative barriers in the health system during linkage to care, which cumulatively resulted in some of them disengaging from care due to frustration or resignation. By also capturing HCWs’ perspectives, we were able to contextualize these health system barriers to show that they were not simply due to ineptitude but rather the result of deficits in human resources or in the material support HCWs needed to effectively do their jobs.

For example, difficulties in navigating within and between health facilities—especially between DMCs where patients got diagnosed and DOT centers where treatment was initiated—was the most common barrier reported by patients. Numerous studies and a recent systematic review have highlighted the circuitous pathways that patients traverse before getting diagnosed and starting on TB treatment in India,^25-28^ especially for those who initially seek care in the private sector.^25 27^ Our study reveals similarly circuitous pathways within public sector care that contributed to some patients not starting treatment, despite getting diagnosed. Patients were often not provided with the required information to make it to a DOT center and start treatment. Some patients in our sample had not returned to find out their sputum microscopy result and had not been notified of the positive result by phone.

From the perspective of HCWs, navigational challenges faced by patients were the result of constraints HCWs faced, such as lack of material support for making phone calls to patients or HCWs at other facilities. These constraints prevented HCWs from notifying patients of their TB diagnoses or coordinating transfer of care between facilities. HCWs also did not have administrative tools, such as information on addresses of DOT centers, to provide patients with accurate referral information. As such, these navigational challenges have the potential to be addressed through specific interventions to improve communication and coordination, including short messaging service (SMS) notifications to patients about test results,^29 30^ electronic health records with real-time availability of patient information across facilities,^20 30^ paper-based or electronic directories with detailed information to facilitate referral to other facilities, or employment of patient navigators (individuals designated to help patients traverse the health system).^31^ Similarly, other barriers in pathway 1 are potentially rectifiable through practical changes to the health system, including hiring of more personnel, providing greater material support to HCWs (for phone calls, etc.), and reducing administrative hurdles for patients.

The second pathway suggested that negative and judgmental HCW attitudes regarding patients provoked patient attitudes of distrust towards the health system, ultimately resulting in refusal of further care by either the patient or the health system. HCWs conducted some activities with good intentions, such as home visits or disclosing a patient’s diagnosis to community leaders; however, the manner in which such activities were implemented were often perceived by patients to be a violation of autonomy. In other cases, HCWs labeled patients as being “disobedient” or “irregular,” judgments that resulted in negative consequences for these patients, such as delayed registration or frank refusal of care by the health system. Even when such TB patients were not refused further care by HCWs, patients felt devalued, resulting in their own refusal of further care.

Our prior quantitative study was one of the first to show that patients with a prior TB treatment history have a higher PTLFU rate than new TB patients,^18^ which is of concern as these patients are more likely to have drug-resistant TB. In the current qualitative study, delayed registration and refusal of care by HCWs were particular problems for patients with a prior treatment history.^32^ Whether intentional or not, delayed registration of these patients by HCWs may result in the appearance of more favorable programmatic outcomes (e.g., higher rates of TB treatment completion).^18^ Such actions by HCWs not only contribute to a lack of transparency and accountability by the health system, but they may also result in exclusion of the most socially marginalized patients from care.^33^

A key implication of this second pathway is that, because critical tensions lie within the HCW-patient interaction, addressing this problem may require sensitization of HCWs to transform their attitudes and behaviors. In addition, ongoing tensions in TB patients’ interactions with HCWs could continue to contribute to patient loss to follow-up during the TB treatment course, as has been suggested in prior literature from India^34 35^ and globally.^16^ Our findings support the importance of systematically measuring and improving patients’ user experience of the health system.^36^ Assessing user experience through periodic patient surveys and incentivizing HCWs who receive good reports may be another approach for improving the HCW-patient interaction. Our findings are consistent with prior qualitative studies highlighting considerable patient hardship within India’s facility-based DOT model,^35^ although this model is currently changing in parts of the country with introduction of digital adherence technologies that have facilitated self-administered therapy for some patients.^37 38^

Although our explanatory model focuses on barriers within the health system, our intention is not to minimize patient-, family-, and society-related barriers contributing to PTLFU, which were described in thematic category 1 (Table 1). Alcohol use disorder was particularly common and has been previously described as a barrier to TB care in Chennai, especially in patients with a prior treatment history.^39^ Interventions to address alcohol use have been associated with promising improvements in treatment success.^40^ Family- and society-related barriers also included lack of social support and TB-related stigma—which were particularly a problem for women who had difficulties traveling to health facilities if not accompanied by a family member. Some patients had advanced TB disease at the time of diagnosis, which made it more difficult to navigate the health system and also resulted in some patients dying before official registration. This highlights a need to diagnose patients earlier in the disease course, by reducing diagnostic delays through active case-finding initiatives^41^ and better training of HCWs, who frequently “miss” patients presenting with early TB symptoms.^42 43^ Challenges at the level of patients, their families, or society were often amplified by the health system barriers described in our explanatory model. For example, alcohol use disorder often contributed to moral judgments of patients being “disobedient” by HCWs, and advanced illness accentuated the already considerable challenges of navigating the health system.

Strengths of our study include its in-depth analysis of narrative data from patients and HCWs to assemble an explanatory model that may provide an integrated framework for addressing health system problems contributing to PTLFU. In addition, qualitative data collection was embedded within a quantitative cohort study, allowing us to achieve a more nuanced understanding of PTLFU than is possible using either approach alone. For example, after finding that patients with a prior treatment history have greater risk of PTLFU,^18^ we delved into the unique challenges faced by these patients in the qualitative study. The main limitation of this study is that more than one-third of PTLFU patients were untrackable by researchers due to missing contact information in government records, and a few patients declined to participate in the qualitative interviews.

## Conclusions

In this qualitative study, we found that PTLFU is a complex problem involving a diverse reasons that TB patients do not start treatment or get officially registered in the TB program. A unifying finding of our explanatory model was the prominent role of health system-related barriers in contributing to PTLFU for nearly all patients—both directly and by amplifying patient-related challenges to engaging in care. Health system barriers play a particularly prominent role for patients with a prior TB treatment history, who are at higher risk for having drug-resistant TB. Addressing this gap in TB care delivery will therefore require development of theory-informed multi-faceted interventions that address organizational and administrative health system barriers and transform the attitudes of HCWs towards TB patients.^44^ Such interventions should focus on making it easier for patients to navigate the health system, ensuring prompt communication of TB diagnoses to patients, reducing constraints that prevent HCWs from effectively performing their jobs, and educating HCWs in non-judgmental approaches to TB care. These strategies have the potential to reduce PTLFU while ensuring patient-centered care to all individuals living with TB, as has been envisioned by the World Health Organization’s End TB Strategy.^45^

## Data Availability

The qualitative data have not been included as a supplement to this manuscript because this would likely compromise the individual privacy of patients, as it may be possible to identify specific individuals based on the in-depth manuscripts. Requests for the de-identified qualitative dataset can be made by contacting Dr. Beena Thomas, although access to these data may be subject to review by the NIRT Institutional Ethics Committee.

## List of Abbreviations

CI: confidence interval
DMC: designated microscopy center
DOT: directly observed therapy
FGD: focus group discussion
HCW: healthcare worker
HV: health visitor
LT: laboratory technician
PTLFU: pretreatment loss to follow-up
RNTCP: Revised National Tuberculosis Control Programme
STS: senior treatment supervisor
TB: tuberculosis

## Ethics approval and consent to participate

The research protocol for this study was approved by the Institutional Ethics Committee of the National Institute for Research in TB (NIRT) (FWA00005104) on December 29, 2014 and the Institutional Review Board of Brigham and Women’s Hospital (Partners Healthcare) (FWA00000484) on January 13, 2015. Written informed consent was collected from all PTLFU patients who agreed to participate in the qualitative interviews. Note that, in addition to anonymizing patient names and other identifying information, in many cases we have anonymized names of health facilities if patient or family member comments might reflect poorly upon that health facility.

## Consent for publication

Quantitative data were de-identified prior to analysis, and care has been taken to ensure that specific patients or health facilities cannot be identified based on the narrative excerpts included in the manuscript.

## Authors’ contributions

BET led project management and supervision of field data collection, and she supported study conceptualization, design, and data interpretation. S Sellappan contributed to study design, project management, and field data collection. CS contributed to project management and field data collection. JL contributed to study conceptualization, study design, and facilitated field data collection. SL contributed to field data collection and data entry. AR contributed to field data collection and data entry. BJ contributed to field data collection and data entry. SK contributed to field data collection and data entry. SA contributed to field data collection and data entry. ML contributed to data analysis, data interpretation, and initial drafting of the manuscript. ATG contributed to data analysis, data interpretation, and initial drafting of the manuscript. KM contributed to study conceptualization, study design, acquisition of funding, and data interpretation. S Swaminathan contributed to study conceptualization, study design, and data interpretation. RS led study conceptualization, study design, acquisition of funding, data analysis, data interpretation, and initial drafting of the manuscript. RS also contributed to data entry, project management, and supervision of field data collection. All authors provided critical revisions to the initial manuscript draft and approved the final paper.

## References

1. Subbaraman R, Nathavitharana RR, Mayer KH, et al. Constructing care cascades for active tuberculosis: A strategy for program monitoring and identifying gaps in quality of care. PLoS Med 2019;16(2):e1002754.

2. MacPherson P, Houben R, Glynn JR, et al. Pre-treatment loss to follow-up in tuberculosis patients in low- and lower-middle-income countries and high-burden countries: a systematic review and meta-analysis. Bull World Health Organ 2014;92(2):126–38.

3. Naidoo P, Theron G, Rangaka MX, et al. The South African Tuberculosis Care Cascade: Estimated Losses and Methodological Challenges. J Infect Dis 2017;216(suppl_7):S702–S13.

4. Subbaraman R, Nathavitharana RR, Satyanarayana S, et al. The Tuberculosis Cascade of Care in India’s Public Sector: A Systematic Review and Meta-analysis. PLoS Med 2016;13(10):e1002149.

5. Dandona R, Dandona L, Mishra A, et al. Utilization of and barriers to public sector tuberculosis services in India. Natl Med J India 2004;17(6):292–9.

6. Gopi PG, Chandrasekaran V, Subramani R, et al. Failure to initiate treatment for tuberculosis patients diagnosed in a community survey and at health facilities under a DOTS program in a district of south India. Indian J Tuberc 2005;52:153–56.

7. Mehra D, Kaushik RM, Kaushik R, et al. Initial default among sputum-positive pulmonary TB patients at a referral hospital in Uttarakhand, India. Trans R Soc Trop Med Hyg 2013;107(9):558–65.

8. Pillai D, Purty A, Prabakaran S, et al. Initial default among tuberculosis patients diagnosed in select medical colleges of Puducherry: issues and possible interventions. Int J Med Sci Public Health 2015;4(7):1–4.

9. Mandal A, Basu M, Das P, et al. Magnitude and reasons of initial default among new sputum positive cases of pulmonary tuberculosis under RNTCP in a district of West Bengal, India. South East Asia J of Public Health 2015;4(1):41–47.

10. Tripathy JP, Srinath S, Naidoo P, et al. Is physical access an impediment to tuberculosis diagnosis and treatment? A study from a rural district in North India. Public Health Action 2013;3(3):235–9.

11. Dave P, Nimavat P, Shah A, et al. Knowing more about initial default among diagnosed sputum smear-positive pulmonary tuberculosis patients in Gujarat, India [Abstract PC-868-03]. Int J Tuberc Lung Dis 2013;17 (Suppl 2)(12):S469.

12. Sai Babu B, Satyanarayana AV, Venkateshwaralu G, et al. Initial default among diagnosed sputum smear-positive pulmonary tuberculosis patients in Andhra Pradesh, India. Int J Tuberc Lung Dis 2008;12(9):1055–8.

13. Ahmed S, Autrey J, Katz IT, et al. Why do people living with HIV not initiate treatment? A systematic review of qualitative evidence from low- and middle-income countries. Soc Sci Med 2018;213:72–84.

14. Ware NC, Wyatt MA, Geng EH, et al. Toward an understanding of disengagement from HIV treatment and care in sub-Saharan Africa: a qualitative study. PLoS Med 2013;10(1):e1001369. discussion e69.

15. Merten S, Kenter E, McKenzie O, et al. Patient-reported barriers and drivers of adherence to antiretrovirals in sub-Saharan Africa: a meta-ethnography. Trop Med Int Health 2010;15 Suppl 1:16–33.

16. Munro SA, Lewin SA, Smith HJ, et al. Patient adherence to tuberculosis treatment: a systematic review of qualitative research. PLoS Med 2007;4(7):e238.

17. Daftary A, Padayatchi N, O’Donnell M. Preferential adherence to antiretroviral therapy over tuberculosis treatment: a qualitative study of drug-resistant TB/HIV co-infected patients in South Africa. Glob Public Health 2014;9(9):1107–16.

18. Thomas BE, Subbaraman R, Sellappan S, et al. Pretreatment loss to follow-up of tuberculosis patients in Chennai, India: a cohort study with implications for health systems strengthening. BMC Infect Dis 2018;18(1):142.

19. Dhanaraj B, Papanna MK, Adinarayanan S, et al. Prevalence and risk factors for adult pulmonary tuberculosis in a metropolitan city of South India. PLoS One 2015;10(4):e0124260.

20. Subbaraman R, Thomas BE, Sellappan S, et al. Tuberculosis patients in an Indian mega-city: Where do they live and where are they diagnosed? PLoS One 2017;12(8):e0183240.

21. Creswell J, Klassen A, Clark V, et al. Best practices for mixed methods research in the health sciences: NIH Office of Behavioral and Social Sciences Research, 2011.

22. Creswell JW. Research design: Qualitative, quantitative, and mixed methods approaches. Thousand Oaks, CA: Sage Publications 2013.

23. Noblit G, Hare R. Meta-ethnography: Synthesizing qualitative studies. Newbury Park, CA: Sage 1988:88.

24. Thomas J, Harden A. Methods for the thematic synthesis of qualitative research in systematic reviews. BMC Med Res Methodol 2008;8:45.

25. Sreeramareddy CT, Qin ZZ, Satyanarayana S, et al. Delays in diagnosis and treatment of pulmonary tuberculosis in India: a systematic review. Int J Tuberc Lung Dis 2014;18(3):255–66.

26. Mistry N, Rangan S, Dholakia Y, et al. Durations and Delays in Care Seeking, Diagnosis and Treatment Initiation in Uncomplicated Pulmonary Tuberculosis Patients in Mumbai, India. PLoS One 2016;11(3):e0152287.

27. Veesa KS, John KR, Moonan PK, et al. Diagnostic pathways and direct medical costs incurred by new adult pulmonary tuberculosis patients prior to anti-tuberculosis treatment - Tamil Nadu, India. PLoS One 2018;13(2):e0191591.

28. Yellappa V, Lefevre P, Battaglioli T, et al. Patients pathways to tuberculosis diagnosis and treatment in a fragmented health system: a qualitative study from a south Indian district. BMC Public Health 2017;17(1):635.

29. Wagstaff A, van Doorslaer E, Burger R. SMS nudges as a tool to reduce tuberculosis treatment delay and pretreatment loss to follow-up. A randomized controlled trial. PLoS One 2019;14(6):e0218527.

30. Mehta K, Kumar AMV, Chawla S, et al. ‘M-TRACK’ (mobile phone reminders and electronic tracking tool) cuts the risk of pre-treatment loss to follow-up by 80% among people living with HIV under programme settings: a mixed-methods study from Gujarat, India. Glob Health Action 2018;11(1):1438239.

31. McBrien KA, Ivers N, Barnieh L, et al. Patient navigators for people with chronic disease: A systematic review. PLoS One 2018;13(2):e0191980.

32. Ministry of Health and Family Welfare. Report of the first national anti-tuberculosis drug resistance survey of India 2014-16. New Delhi, India: Ministry of Health and Family Welfare, 2018.

33. Singh V, Jaiswal A, Porter JD, et al. TB control, poverty, and vulnerability in Delhi, India. Trop Med Int Health 2002;7(8):693–700.

34. Jaiswal A, Singh V, Ogden JA, et al. Adherence to tuberculosis treatment: lessons from the urban setting of Delhi, India. Trop Med Int Health 2003;8(7):625–33.

35. Yellappa V, Lefevre P, Battaglioli T, et al. Coping with tuberculosis and directly observed treatment: a qualitative study among patients from South India. BMC Health Serv Res 2016;16:283.

36. Kruk ME, Gage AD, Arsenault C, et al. High-quality health systems in the Sustainable Development Goals era: time for a revolution. Lancet Glob Health 2018;6(11):e1196–e252.

37. Cross A, Gupta N, Liu B, et al. 99DOTS: A low-cost approach to monitoring and improving medication adherence. ICTD ‘19; 2019 January 4-7 2019; Ahmedabad, India.

38. Subbaraman R, de Mondesert L, Musiimenta A, et al. Digital adherence technologies for the management of tuberculosis therapy: mapping the landscape and research priorities. BMJ Glob Health 2018;3:e001018.

39. Suhadev M, Thomas BE, Raja Sakthivel M, et al. Alcohol use disorders (AUD) among tuberculosis patients: a study from Chennai, South India. PLoS One 2011;6(5):e19485.

40. Thomas B, Watson B, Senthil EK, et al. Alcohol intervention strategy among tuberculosis patients: a pilot study from South India. Int J Tuberc Lung Dis 2017;21(8):947–52.

41. Prasad BM, Satyanarayana S, Chadha SS, et al. Experience of active tuberculosis case finding in nearly 5 million households in India. Public Health Action 2016;6(1):15–8.

42. Kwan A, Daniels B, Saria V, et al. Variations in the quality of tuberculosis care in urban India: A cross-sectional, standardized patient study in two cities. PLoS Med 2018;15(9):e1002653.

43. Satyanarayana S, Subbaraman R, Shete P, et al. Quality of tuberculosis care in India: a systematic review. Int J Tuberc Lung Dis 2015;19(7):751–63.

44. Chaisson LH, Katamba A, Haguma P, et al. Theory-Informed Interventions to Improve the Quality of Tuberculosis Evaluation at Ugandan Health Centers: A Quasi-Experimental Study. PLoS One 2015;10(7):e0132573.

45. Uplekar M, Weil D, Lonnroth K, et al. WHO’s new End TB strategy. Lancet 2015;385(9979):1799–801.

